# Divergent Recovery Trajectories: The Influence of Injury Mechanism on Stair-Climbing Outcomes After Traumatic Brain Injury - a TBI Model Systems Study

**DOI:** 10.64898/2026.07.04.26357287

**Authors:** Matthew J. Beth, Jennifer Marwitz, Nojan Valadi, Niyati Baweja, Harsimran S Baweja

## Abstract

**Background/Objectives:** Traumatic Brain Injuries (TBIs) often cause profound functional impairments, yet the influence of TBI mechanisms on stair-climbing functional independence over extended timelines remains poorly understood. This study assesses whether Rasch-transformed FIM Stairs scores varied by TBI mechanism over follow-ups spanning 10 years or more.

**Methods:** Data from the TBI Model Systems database were analyzed. The original 30,768 data entries were reduced to 6,226, corresponding to individuals with at least 10 years of data. Functional Independence Measure Stairs data were transformed to logit units via Rasch analysis before being evaluated with a linear mixed-effects regression, incorporating TBI mechanisms, age, follow-up time, and their interactions, with random effects accounting for the participant ID and pre-injury residence location.

**Results:** TBI mechanisms meaningfully shape very long-term stair-climbing. Gunshot wounds and pedestrian-related accidents are associated with poorer performances, whereas motorcycles, bicycles, unclassified vehicular accidents, winter sports, other sports, and fall-related TBIs demonstrated relatively better function. Age, follow-up time, and their interaction also reached significance.

**Conclusions:** Stair-climbing recovery trajectories over extended time significantly vary by TBI mechanism, with individuals with TBIs from gunshots and pedestrian-related accidents showing the most unfavorable recoveries. These findings support the development of mechanism-specific prognostic guidance and individualized rehabilitation strategies, thereby encouraging tailored approaches to improve outcomes.

## INTRODUCTION

Traumatic Brain Injury (TBI) is a major global health concern, and it is estimated that there are over 50 million new cases annually worldwide (1). Although TBIs are common, only a small proportion are fatal; approximately 90% of affected individuals survive their injuries (2), meaning that approximately 45 million people each year survive their TBI and live with residual disabilities and impairments. As of 2021, global burden estimates indicate that TBIs accounted for approximately 5.48 million years lived with disability worldwide, highlighting the substantial long-term impact of TBIs on population health (3,4). Survivors of TBI experience chronic sequelae that may include impairments in motor functioning, cognition, and emotional regulation. In addition to clinical implications, TBIs impose a significant financial burden on affected individuals and healthcare systems worldwide. TBIs are responsible for roughly 1.5 million deaths each year, along with millions of hospital admissions. Moderate-to-severe TBIs (msTBIs) disproportionately contribute to fatalities, long-term impairment, and healthcare expenditures (5). Ongoing healthcare needs from TBI significantly contribute to global medical spending. The annual global costs exceed 400 billion USD (6). A single severe TBI alone may result in in-hospital costs ranging from approximately 2,130 to over 400,000 USD, with expenses escalating further from long-term rehabilitation, outpatient care, and accommodations to adapt to newly acquired disabilities (7).

Many post-TBI adaptations are necessary to compensate for impaired independence in functional activities (8). In addition to the high medical costs associated with TBIs, reduced functional independence is associated with poorer long-term quality of life (9) and imposes substantial emotional, financial, and social strain on families and society (10,11). Therefore, understanding and addressing compromised functional independence following TBI is a clinical priority and a critical research area.

### Falls

A robust relationship has been documented between TBI mechanisms and long-term functional recovery. Specifically, individuals with TBIs caused by falls tend to demonstrate consistently poorer long-term outcomes relative to those injured through other mechanisms (12,13). Individuals with TBIs resulting from road traffic incidents and sports-related activities are associated with better outcomes than those caused by falls (12). Subsequent research expanded on these findings, demonstrating that individuals with TBIs from traffic accidents frequently sustain the most severe injuries, yet are associated with relatively more favorable outcomes (14). Conversely, fall-related TBIs tend to produce less severe initial injuries while being associated with worse long-term outcomes (14).

### Injury Severity

Latent trajectory modeling has delineated distinct low, moderate, and high functional recovery patterns for the cognitive and motor subscales of the Functional Independence Measure (FIM). Subsequent multinomial logistic regression analyses have estimated odds of class membership based on injury-related characteristics. Findings from the work indicate that penetrating or open head injuries, as well as longer delays between injury occurrence and admission to inpatient rehabilitation, are associated with less favorable motor recovery trajectories (15,16).

Longitudinal research following individuals with TBI for more than 2 decades has demonstrated that substantial functional limitations often persist well into adulthood, even many years after injury. These studies consistently show that both injury severity and injury mechanism are significantly related to enduring disability (13,17). Additional evidence from cohort studies indicates that a considerable proportion of individuals continue to experience motor impairments years after injury, including balance problems and movement-related difficulties affecting the trunk or extremities (18). Across studies, injury severity is negatively associated with long-term functional status, suggesting that individuals with more severe TBIs have worse outcomes. Vocational outcomes are similarly affected, with fewer than half of individuals returning to competitive employment, and many of those who do report reduced earnings attributable to ongoing cognitive and physical limitations secondary to TBI (19).

Person-centered analytic approaches, such as latent profile analysis, have further identified multiple subgroups characterized by progressively greater levels of functional impairment. Common between the analyses was that the most impaired classes were composed of individuals with the most severe TBIs and were characterized by profound cognitive deficits, impaired consciousness, and poor functional independence. Membership in the subgroup is associated with worse functional recovery trajectories and prolonged hospitalizations (11,20).

Complementing these findings, meta-analytic evidence examining outcomes within the first year post-injury has demonstrated a moderate overall association between injury severity and functional outcome, consistent with conventional benchmarks for medium effect sizes. These analyses report substantial heterogeneity across studies, as marked by elevated Cochran’s Q and I² statistics, underscoring variability in outcome estimates. Nevertheless, the aggregated evidence supports injury severity as a robust predictor of functional outcomes during the first year following TBI (21).

### Injury Severity from Falls & Violence

A retrospective analysis demonstrated a clear relationship between TBI severity and the TBI mechanism, with falls from height and firearm injuries disproportionately associated with severe presentations (22). Penetrating injuries from gunshot wounds and other open-head injuries represented more than half of the analysis’s sample (22). Long-term follow-up studies indicate that individuals with msTBIs frequently exhibit substantial neuromotor dysfunction, including chronic gait impairment, balance instability, postural deficits, spasticity, poor limb coordination, and diminished fine motor skills, which can persist for years after the injury (18,23). Furthermore, TBIs caused by falls from height and violent mechanisms are associated with an elevated risk of intracranial hemorrhage, skull fractures, cerebrospinal fluid leaks, and secondary complications, all indicating greater injury severity (11,24).

Taken together, the findings indicate that greater TBI severity is commonly associated with falls and violent mechanisms. Accordingly, the injury severity results can reasonably be generalized to TBIs caused by falls, gunshot wounds, assaults, and other violent TBI mechanisms.

### Traumatic Brain Injury Model Systems

The data analyzed in this study were drawn from the Traumatic Brain Injury Model Systems national database, housed at Craig Hospital in Englewood, Colorado. Established in 1987, the TBIMS was designed to support research, guide quality improvement efforts, and inform evidence-based clinical practice in TBI rehabilitation (25). The TBIMS has been widely recognized as the largest and most comprehensive longitudinal database of individuals with TBI worldwide (26). Currently, the database includes information on more than 20,000 individuals with TBI, collected over nearly 4 decades (27).

The TBIMS gathers data from a broad range of outcome measures, including the Disability Rating Scale (28,29), the Functional Independence Measure (30,31), substance use, arrests, and psychiatric conditions in the previous year. Additionally, the TBIMS collects data on rehospitalization history and measures of social participation and global outcomes derived from the Glasgow Outcome Scale–Extended (32,33), the Supervision Rating Scale (34), the PART-O participation measure (35), and the Satisfaction with Life Scale (36). Data are obtained at rehabilitation admission and discharge, and follow-up data are collected via telephone interviews at 1, 2, 5, and 10 years post-injury, and at 5-year intervals thereafter (37).

Within the TBIMS, the mechanism of injury is documented using the Cause variable. The nominal variable includes 19 distinct levels, including motor vehicle accidents, motorcycle injuries, bicycle injuries, all-terrain vehicle incidents, other unclassified vehicular injuries, gunshot wounds, assaults with a blunt instrument, other violent mechanisms, water sports, track and field sports, gymnastic activities, winter sports, air sports, other sports, falls, injuries from being struck by a falling or flying objects, pedestrian injuries, and other unclassified TBI mechanisms.

### Functional Independence Measure

The data analyzed in the study were obtained using the Functional Independence Measure (30,31). The FIM is an 18-item assessment designed to quantify functional independence and the burden of care relating to motor (13 items) and cognitive (5 items) functioning. Items are rated on a 7-point scale, ranging from total assistance (1) to complete independence (7) (38). The comprehensive instrument evaluates multiple functional domains, including problem-solving, memory, expression, comprehension, social interaction, eating, bathing, grooming, upper-body and lower-body dressing, toileting, bowel and bladder management, and transfers (38). At admission and during inpatient rehabilitation, FIM assessments are administered by trained clinicians through direct observation of the functional tasks. Subsequent follow-up data are collected through structured telephone interviews (37).

Application of the FIM in neurotrauma rehabilitation has been extensively examined over several decades. Early evidence in 1998 demonstrated excellent interrater reliability (*r* =.95), with the motor subscale exhibiting the strongest reliability (39). More recently, findings from 2016 indicated that the FIM also demonstrates excellent internal consistency, with a Cronbach’s alpha of *α* =.98 (38).

### Rasch Analysis

FIM scores are prone to pronounced ceiling effects, necessitating the use of Item Response Theory (IRT) methods before statistical analysis. An appropriate IRT framework is provided by a Rasch analysis, which converts ordinal items into interval-level logit estimates using the partial credit model. The transformation is paramount because linear models assume equal distances between scale points. Failure to meet the assumption may distort estimates and bias standard errors (40). For example, a one-unit difference between the raw scores of 3 (“moderate assistance”) and 4 (“minimal assistance”) does not inherently represent the same degree of functional change as another one-unit difference between scores of 5 (“supervision”) and 6 (“modified independence”). Rasch analysis rectifies this limitation by mapping the original 1–7 ordinal responses onto a continuous logit scale in a uniformly spaced manner (41,42). While the original data reflect ordinal FIM Stairs scores, the transformation yields an evaluation of the underlying latent stair-climbing ability. In addition to establishing interval-level logit scores, Rasch analyses improve scale linearity and residual normality, strengthen the accuracy and power of both fixed-and random-effects estimates, and support reliable comparisons across individuals and groups.

Despite evidence that TBI outcomes vary by mechanism of injury, much of the current literature has focused on associations with functional outcomes over shorter timeframes (often 1-8 years). Studies have frequently utilized raw FIM scores, which may mask meaningful differences between groups. While the relationship between the mechanism of injury and short-term functional outcomes is relatively well characterized, a significant gap remains in understanding whether TBI mechanisms are associated with long-term functional recovery trajectories when FIM stairs scores are evaluated with greater precision by transforming the scores into logit scores using Rasch analysis. To address the gap, this study applies a linear mixed-effects regression to examine longitudinal trends in FIM stairs scores across TBI mechanisms.

Specifically, this study investigates whether Rasch-transformed FIM stairs scores differ by TBI mechanisms over follow-up periods exceeding 10 years. We hypothesize that individuals who sustain TBIs from falls or violence-related mechanisms will demonstrate lower Rasch-FIM Stairs scores relative to their scores of those injured through automotive or recreational mechanisms.

## METHODS

### Participants

Data received from the TBIMS database included records for 30,768 individuals. With the focus on evaluating the impact of TBI mechanisms on very long-term stair-climbing functional independence, participants with less than 10 years of available follow-up data were excluded from the analysis. The exclusion reduced the sample size to 6,226 individuals.

### Statistical Analysis

A linear mixed-effects regression model was used to evaluate predictors of stair-climbing functional independence. The model designated the “FIMStairsF” variable as the dependent variable, rather than “FIMStairs,” because “FIMStairsF” represents the longitudinal follow-up data. Two fixed effects were incorporated into the model. The first was the TBI mechanism (Cause), a nominal predictor comprising 19 categories: motor vehicles (*n* = 2875), motorcycles (*n* = 513), bicycles (*n* = 141), all-terrain vehicles/cycles (*n* = 97), unclassified vehicular accidents (*n* = 23), gunshot wounds (*n* = 319), assaults (*n* = 658), other violence (*n* = 44), being struck by falling or flying objects (*n* = 73), falls (*n* = 887), pedestrian-related injuries (*n* = 473), unclassified injuries (*n* = 32), gymnastics (*n* = 1), and water (*n* = 6), track (*n* = 7), winter (*n* = 34), air (*n* = 2), and other sports (*n* = 46). The second fixed effect was an interaction term (AGENoPHIF x DAYStoFUF) that combined participant age (AGENoPHIF) with the number of days until follow-up (DAYStoFUF). This term was included to assess whether longitudinal recovery trajectories differed by participant age.

Random effects were specified to account for hierarchical clustering within the data and included participant identifier (Mod1Id) and pre-injury geographic region of residence (ZipInj). Variance components associated with these random factors were estimated and are presented in Table 4.

Statistical significance of the fixed effects was evaluated using t-tests for individual parameter estimates, while F-tests were used to assess the significance of the Cause variable and the interaction term. The alpha level was set at 0.05 (43). All statistical analyses were performed using R (v4.5.1) (44). The Rasch analysis was conducted using the’TAM’ R package (45), and descriptive statistics for the FIM Stairs item, reported in Table 1 for both raw and Rasch-transformed scores, were computed using the mosaic R package (46). After transforming the data into interval-level logit scores via Rasch analysis, a linear mixed-effects regression model was fit to examine the data. Model estimation was performed using the lme4 R package (47), and graphical representations were generated using the ggplot2 R package (48).

**Table 1.**
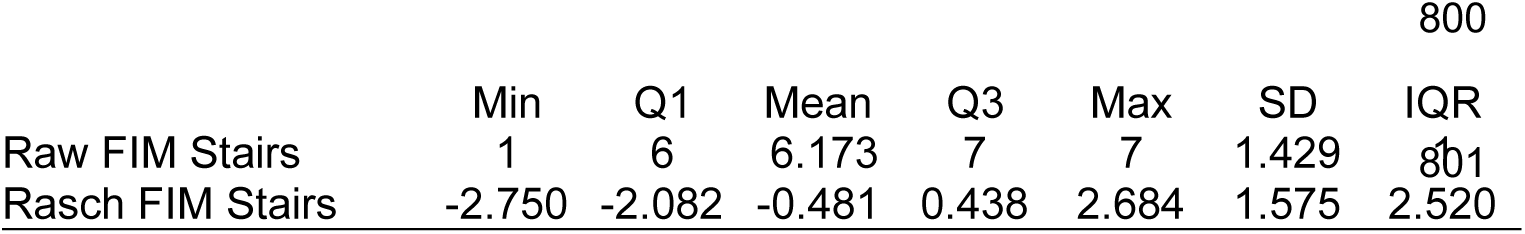
Descriptive statistics of the raw FIM Stairs scores and the Rasch-transformed logit FIM Stairs scores.

### Very Long-Term

Although no consensus definition has been established, the phrase “very long-term” is commonly used in the literature to characterize follow-up data collection over periods that exceed what is considered long-term (49,50). Many studies rely on data collected over 10 years or less, but the TBIMS dataset spans nearly 4 decades, underscoring its comprehensive scope. Therefore, the study defines “very long-term” as follow-up data spanning 10 years or more, aiming to inspire confidence in its extensive data coverage.

## RESULTS

The linear mixed-effects model demonstrated that TBI mechanisms, age at injury, follow-up timing, and their interaction significantly influenced long-term stair-climbing outcomes, highlighting the importance of these factors in recovery trajectories.

Several TBI mechanisms were significantly associated with very long-term stair-climbing functional performance. Individuals with TBIs from motorcycles (Estimate = 0.129, SE = 0.049, *t* = 2.66, *p* =.008), bicycles (Estimate = 0.319, SE = 0.091, *t* = 3.50, *p* <.001), unclassified vehicular accidents (Estimate = 0.660, SE = 0.212, *t* = 3.12, *p* =.002), winter sports (Estimate = 0.517, SE = 0.163, *t* = 3.17, *p* =.002), other sports (Estimate = 0.334, SE = 0.146, *t* = 2.29, *p* =.022), and falls (Estimate = 0.086, SE = 0.042, *t* = 2.06, *p* =.039) were positively associated with very long-term stair-climbing functional ability. Conversely, gunshot wounds (Estimate = −0.448, SE = 0.064*, t* = −6.95, *p* <.001) and pedestrian-related TBIs (Estimate = −0.149, SE = 0.054, *t* = −2.76, *p* =.006) corresponded to significantly poorer stair-climbing functional performance. TBIs from MVAs, ATVs/ATCs, assaults, other violence, water sports, track and field, gymnastics, air sports, unclassified injury mechanisms, and unknown injury origins did not reach statistical significance. Complete estimates, standard errors, t-values, and p-values for all TBI mechanisms are reported in Table 2.

**Table 2.** Summary and coefficient estimates from the fixed effect Cause variable in the linear mixed-effects regression with the Rasch-FIM Locomotion scores as the outcome variable.

| TBI Mechanism | Estimate | SE<sup>1</sup> | t-value | p-value |
| --- | --- | --- | --- | --- |
| MVAs<sup>2</sup> | -0.644 | 1.427 | -0.451 | 0.652 |
| Motorcycle | 0.129 | 0.049 | 2.659 | 0.008** |
| Bicycle | 0.319 | 0.091 | 3.501 | <0.001*** |
| ATV/ATC<sup>3</sup> | 0.155 | 0.102 | 1.524 | 0.128 |
| Unclassified Vehicular | 0.660 | 0.212 | 3.116 | 0.002** |
| Gunshot Wounds | -0.448 | 0.064 | -6.952 | <0.001*** |
| Assaults | 0.067 | 0.050 | 1.354 | 0.176 |
| Other Violence | 0.003 | 0.165 | 0.021 | 0.984 |
| Water Sports | 0.518 | 0.413 | 1.254 | 0.210 |
| Track & Field Sports | 0.835 | 0.476 | 1.756 | 0.079 |
| Gymnastics | 0.635 | 0.964 | 0.659 | 0.510 |
| Winter Sports | 0.517 | 0.163 | 3.169 | 0.002** |
| Air Sports | 0.695 | 0.660 | 1.053 | 0.293 |
| Other Sports | 0.334 | 0.146 | 2.287 | 0.022* |
| Falls | 0.086 | 0.042 | 2.061 | 0.039* |
| Hit By Falling/Flying Object | 0.095 | 0.119 | 0.799 | 0.425 |
| Pedestrian | -0.149 | 0.054 | -2.758 | 0.006** |
| Other Unclassified | -0.070 | 0.188 | -0.372 | 0.710 |
| Unknown | 0.427 | 0.454 | 0.941 | 0.347 |

\*\*\* $p < 0.001$ ; \*\* $0.001$ ; \* $0.01$
<sup>1</sup>Standard Error; <sup>2</sup>Motor Vehicle Accidents; <sup>3</sup>All-Terrain Vehicle/Cycle

Although the age-by-follow-up interaction was statistically significant, the small and inconsistent individual effects suggest a general trend rather than distinct differences across age groups, emphasizing the complexity of recovery over time.

The random-effects structure revealed significant between-participant variability in very long-term stair-climbing functional ability (participant-level *σ*² = 0.569) as well as smaller yet meaningful variability attributed to pre-injury residential zip code (σ² = 0.097), with a residual variance of 1.655. Random effects variance statistics are provided in Table 4. The study’s findings identify TBI mechanisms as a meaningful predictor of very long-term functional recovery in stair climbing. Gunshot wounds and pedestrian-related TBIs were associated with the least favorable outcomes. Meanwhile, several sports-related TBI mechanisms were associated with more favorable recovery trajectories. The significant effects of age, follow-up, and their interaction reinforce the idea that recovery is not uniform across age groups and that recovery is a lifelong journey.

## DISCUSSION

The purpose of this study was to evaluate how TBI mechanisms influence very long-term stair-climbing functional recovery, using Rasch-transformed FIM Stairs scores collected over 10+ years post-TBI. The TBI mechanism emerged as a robust predictor of very long-term stair-climbing functional independence, as evidenced by a significant omnibus effect (F = 6.832, *p* <.0001; see Table 3). Figure 1 depicts TBIs resulting from gunshot wounds and pedestrian-related injuries, which yielded significant negative parameter estimates. An example of a “pedestrian-related accident” would be someone walking along a sidewalk who suffers a TBI by being struck by, or colliding with, a motor vehicle passing by (51). Between the groups, gunshot-related TBIs were associated with consistently lower FIM Stairs scores over the extended follow-up period (see Figure 2)

**Figure 1.**
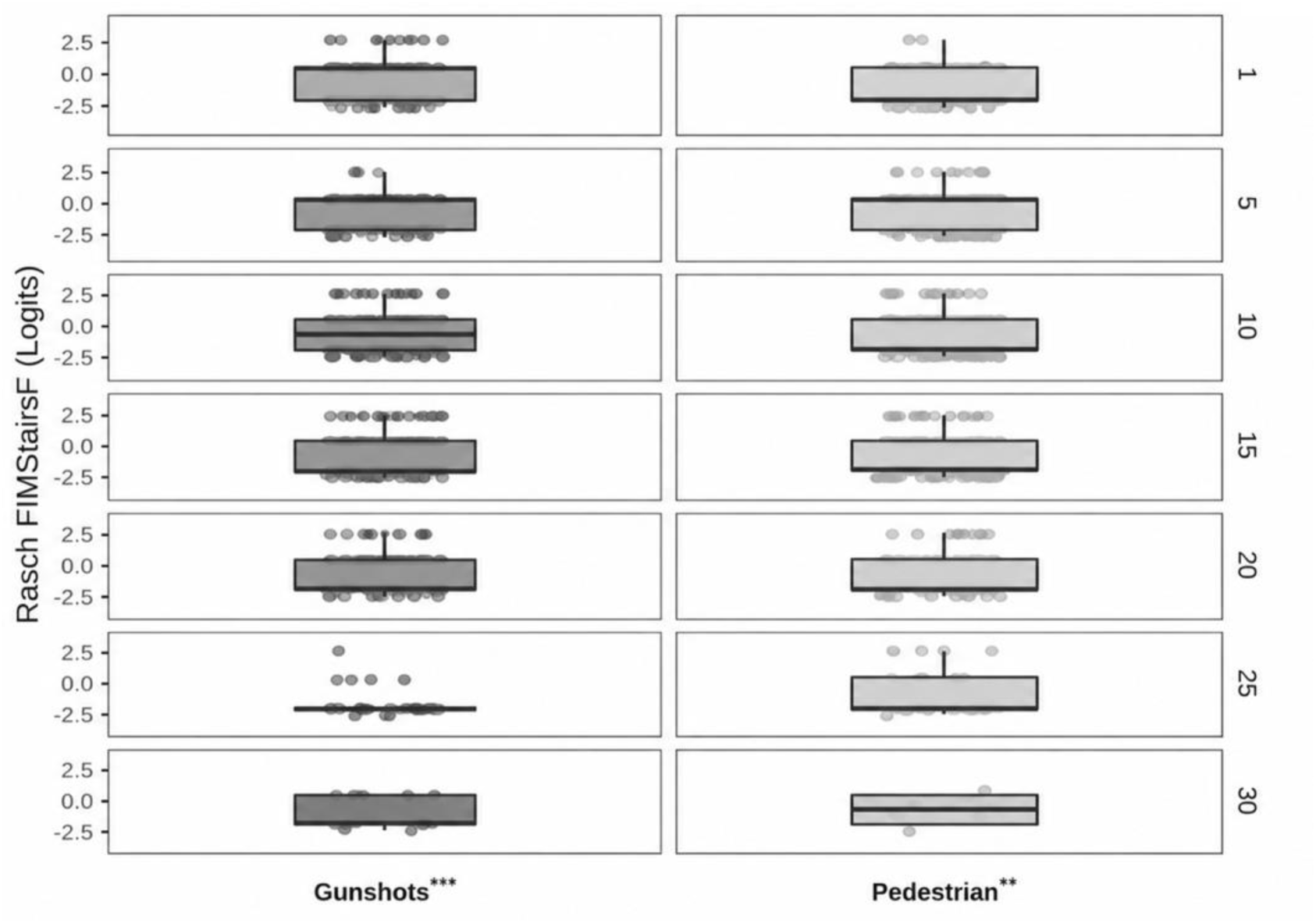
Longitudinal distributions of Rasch-transformed FIM Stairs scores by TBI mechanism. Each column represents individuals stratified by their respective TBI mechanisms, with significant negative estimates (gunshots and pedestrian-related accidents). Each row represents the Rasch-transformed FIM Stairs scores at follow-up years 1, 5, 10, 15, 20, 25, and 30. The box plots depict the median and interquartile range, with individual points representing participant observations. Across follow-ups, violence-related mechanisms (gunshots, assaults, and other violence) demonstrate poorer median stair-climbing abilities, whereas falls, pedestrian injuries, and TBIs from falling or flying objects demonstrate relatively higher levels of functional recovery. Mechanism-related differences in stair-climbing performance persist over extended periods, indicating distinct longitudinal recovery trajectories.

**Figure 2.**
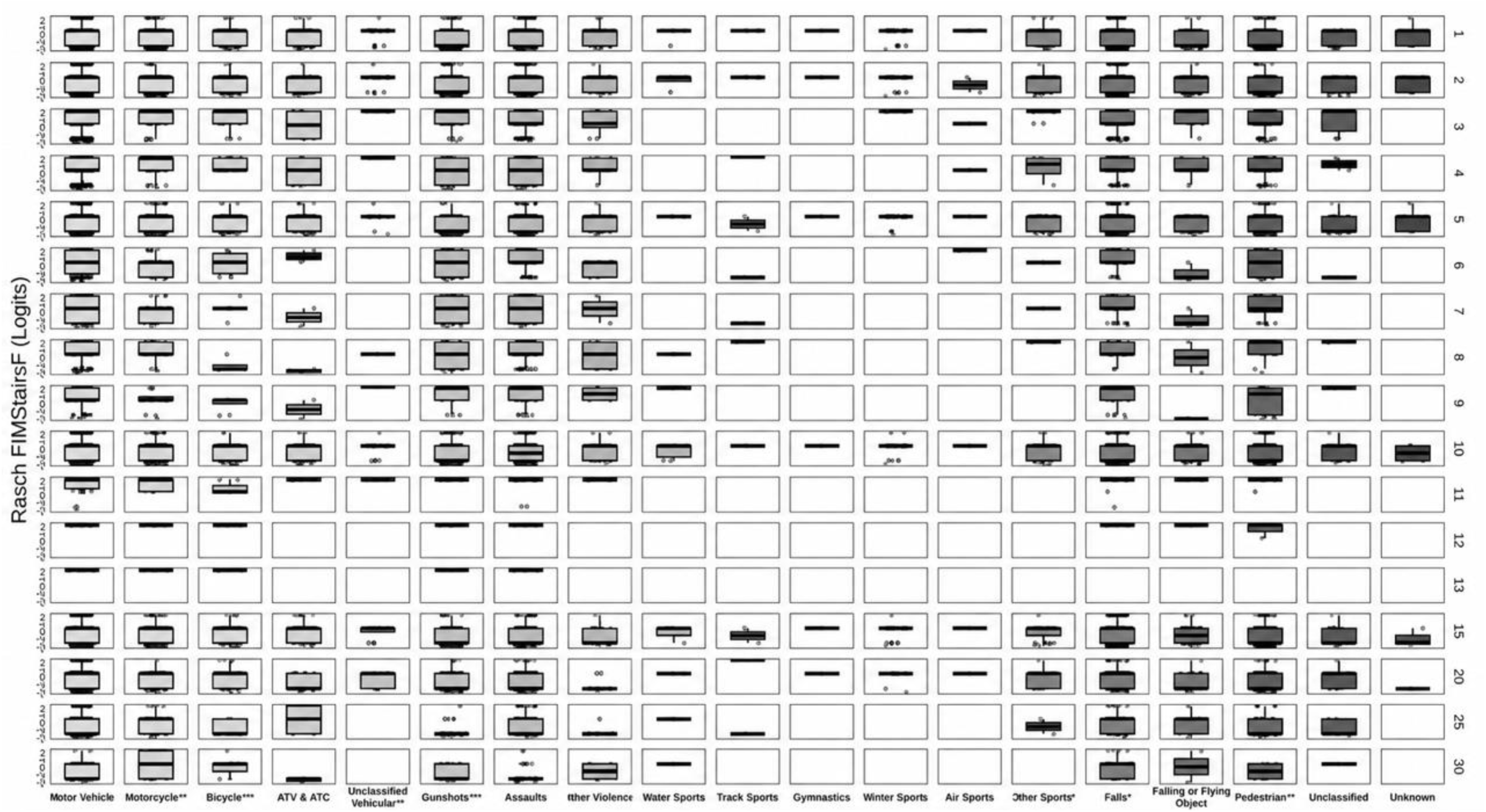
Longitudinal trajectories of Rasch-transformed FIM Stairs scores by TBI mechanism; Each column represents a distinct TBI mechanism, and each row corresponds to the measure at each follow-up period. Sports-related TBI mechanisms show higher and more stable functional trajectories over time, whereas fall-and violence-related mechanisms demonstrate lower median performance and greater variability. Motor vehicle mechanisms exhibit intermediate recovery patterns, with early improvement that eventually plateaus.

**Table 3.**
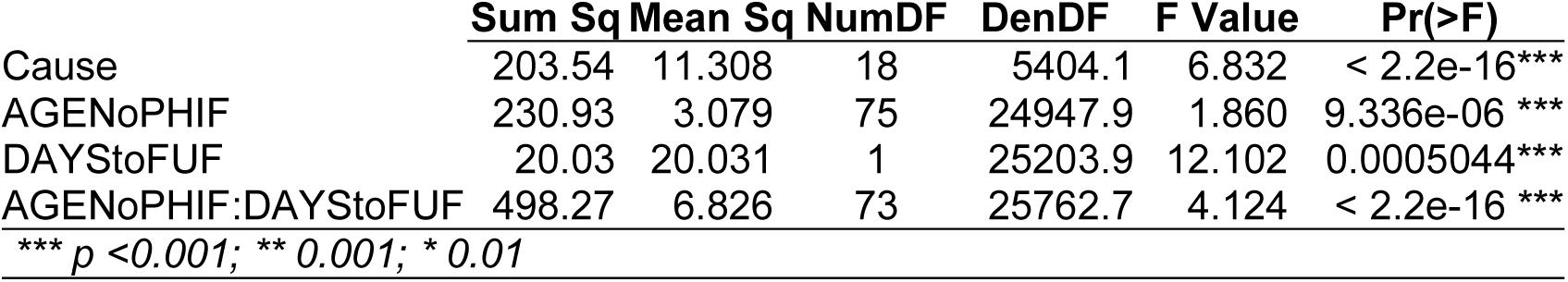
Analysis of Variance (ANOVA) table of the components of the model’s fixed808 effects.

**Table 4.**
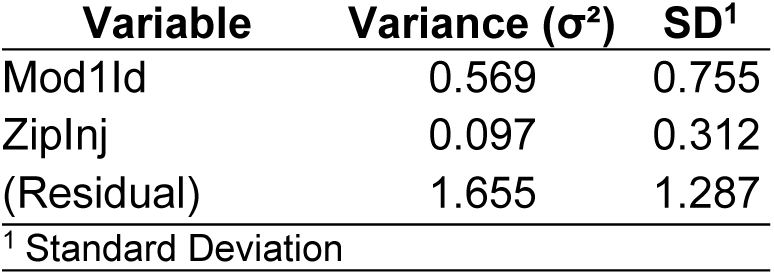
Variance components of the model’s random effects structure (Mod1Id and ZipInj).

Unlike other TBI mechanisms, penetrating TBIs from gunshots uniquely confer prolonged vulnerability to secondary and tertiary health complications well beyond the acute phase of TBIs. When classified into early (acute), intermediate, and late symptom stages, individuals with penetrating TBIs after the acute phase of their injury may develop seizures, refractory cerebral edema, acute hydrocephalus, vasospasm, cerebrospinal fluid leaks, deep venous thrombosis, and traumatic pseudoaneurysms during the intermediate recovery period (53) In later stages, chronic sequelae may include infections, delayed hydrocephalus, cerebrospinal fluid fistulae, venous sinus thrombosis, trephination syndrome, scalp necrosis, arteriovenous fistulas, metal toxicity from retained bullet fragments, temporalis muscle atrophy, and subdural hygromas (54). The association between the long-term medical complications and penetrating TBIs likely significantly contributes to the poorer functional independence observed in individuals with TBIs from gunshot wounds.

Conversely, statistically significant positive estimates were identified for individuals sustaining TBIs caused by motorcycles, winter sports, bicycles, and fallsIndividuals sustaining injuries while engaging in winter sports and bicycle-related activities are likely to bring a more athletic background to their recovery and may have a higher baseline level of physical fitness. Although human-based research is sparse, evidence from animal models suggests that pre-injury exercise may enhance recovery outcomes after TBI. Rodents that participated in voluntary exercise prior to TBI onset demonstrated significantly improved behavioral and functional outcomes post-TBI (55,56). Neurobiological analyses revealed that the brains of physically conditioned animals demonstrate increased expression of neuroprotective and angiogenic factors, including Vascular Endothelial Growth Factor A and EPO (56). The physiological adaptations would aid individuals in a more complete recovery and thereby explain the superior long-term functional outcomes observed in mechanisms that are likely to involve more physically active populations.

TBI mechanisms that did not yield statistically significant estimates included MVAs, all-terrain vehicle/cycle (ATV/ATC) accidents, assaults, other violence, track-and-field sports, water sports, and gymnastics. Within the mixed-effects model, both the participant identifier (Mod1Id) and the pre-TBI geographical location of residence (ZipInj) were statistically significant predictors (see Table 4). Despite this, the residual variance still accounted for the majority of the variance, indicating that a significant proportion of the variability in stair-climbing function remains attributable to individual differences after accounting for both fixed and random effects. The findings partially confirm the study hypothesis: the hypothesis is correct in that a pronounced decline in very long-term stair-climbing ability was observed among individuals with TBIs resulting from gunshot wounds, whereas meaningful improvements in very long-term stair-climbing ability were observed among individuals with TBIs from bicycles, winter, and other sports. However, some study results contradicted the hypothesis. Unexpected findings included significant, long-term functional improvements in stair climbing observed in individuals with TBIs from falls, motorcycle accidents, and unclassified vehicular accidents. It was also unexpected that no significant differences in functional stair-climbing were observed among individuals with TBIs from MVAs, ATVs/ATCs, assaults, other violence, gymnastics, track, air, and water sports. Figure 3 tracks average Rasch-FIMStairsF scores over 30 years for individuals stratified by TBI mechanism, visualizing the study’s hypothesis.

**Figure 3.**
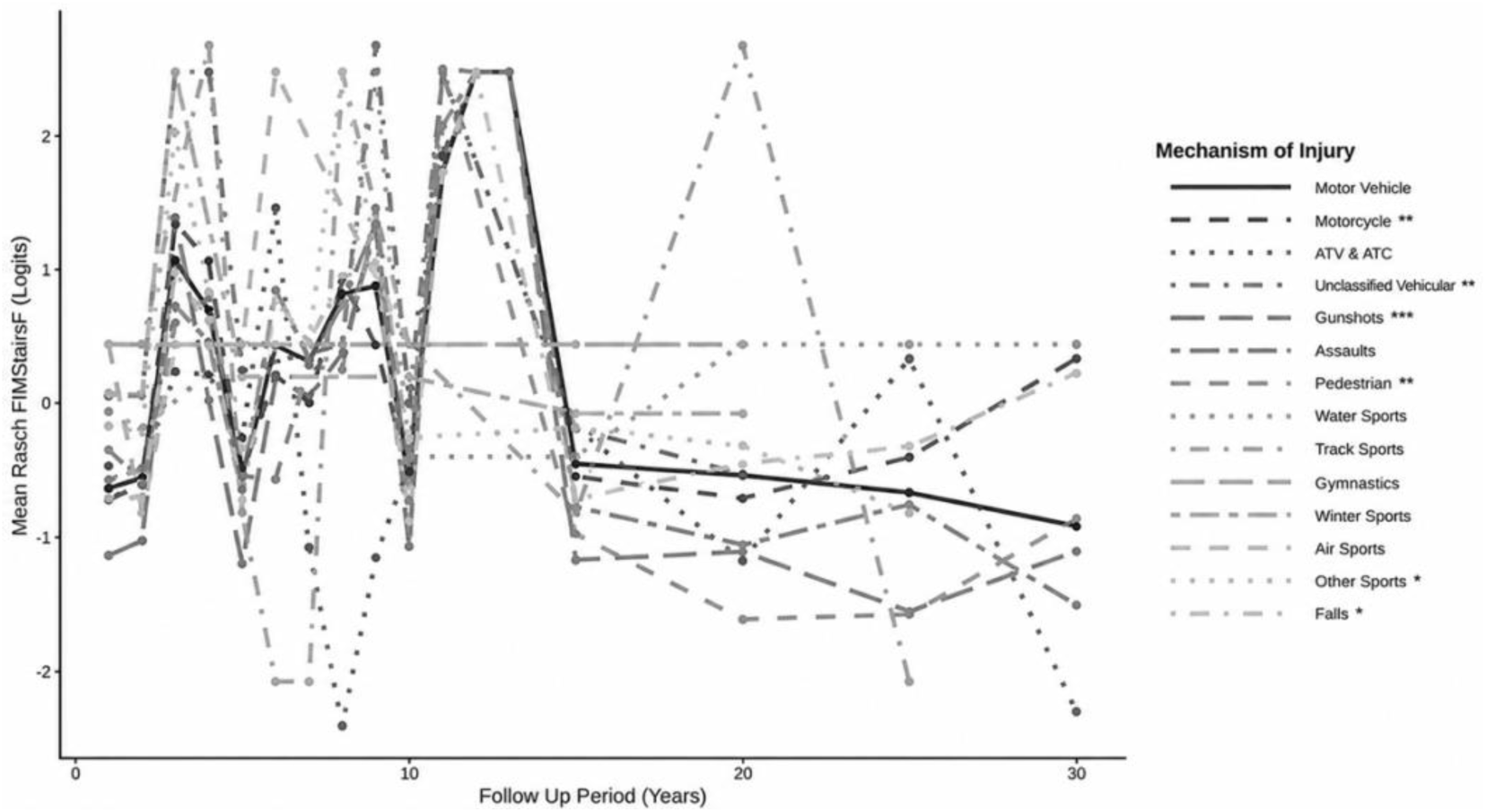
Trajectories of Mean Rasch-FIMStairsF Scores by Mechanism of TBI Across Follow-Up Periods. Mean Rasch-Functional Independence Measure (FIM) Stairs scores (logits) across 30 years for hypothesis-related TBI mechanisms. Each line represents a distinct mechanism of injury, differentiated by line type. Higher logit values indicate greater independence with stair negotiation. The Y-axis represents the mean Rasch-transformed FIMStairsF scores, with logit values from-2.750 to 2.684. Substantial differences across TBI mechanisms are evident in the early years, while later years are characterized by convergence at negative logit values, consistent with lasting limitations in stair performance.

### Rasch Interpretation

Rasch modeling was applied to convert the raw FIM Stairs data into interval-level, logit scores of stair-climbing functional ability. The logit values indicate one’s latent stair-climbing functional capacity, enabling score changes to be interpreted as equal units of functional change across the scale continuum. As such, coefficients from linear mixed-effects models reflect substantive differences in latent functional ability rather than changes in the raw FIM scores. The transformation enhances the suitability of longitudinal analyses and facilitates precise interpretations of recovery trajectories across TBI mechanisms. Within Rasch frameworks, changes of 0.5–0.7 logits are considered as moderate effects (57). The FIM Stairs item is scored on a 1–7 ordinal scale. Following Rasch analysis, logit scores in the study ranged from −2.75 to 2.68 logits (M = −0.48 logits), with an interquartile range of 2.52 logits (Q1 = −2.08 logits, Q3 = 0.44 logits) (see Table 1).

### Age × Follow-Up Interaction

The analysis identified a significant interaction between age at injury (AGENoPHIF) and follow-up time (DAYStoFUF) for predicting very long-term stair-climbing functional independence, F(73, 25,762.7) = 4.12, *p* <.001. The interaction underscores the importance of understanding how age influences recovery, a theme that should resonate with medical professionals and researchers dedicated to improving patient outcomes.

Stair-climbing requires greater physical demands relative to average walking, as it requires more dynamic balance, lower-extremity strength, coordination, and cardiovascular endurance. As such, degeneration of musculoskeletal capacity associated with normative aging will likely be evident in very long-term stair-climbing performance. Recognizing this, rehabilitation specialists can feel encouraged to tailor interventions that address these specific challenges faced by older individuals with TBIS, potentially improving their very long-term mobility.

The analysis confirms that advanced age is associated with impaired long-term functional improvement. Despite the small magnitude of individual interaction estimates, the consistency of estimates across the age spectrum highlights a systematic, age-related influence on recovery trajectories, warranting further investigation into clinical significance.

Clinically, these findings highlight the need to tailor rehabilitation plans to address higher-level mobility functions in aging TBI patients. Recognizing that stair-climbing independence influences community participation and fall risk can motivate professionals to develop more comprehensive, age-sensitive protocols that focus on dynamic balance, strength, and endurance training, thereby improving patient care.

### Recovery Trajectories

A matrix of boxplots showing the logit-FIMStairsF scores for individuals, stratified by TBI mechanism, across follow-up data collections is shown in Figure 2. Each column of boxplots shows the logit-FIMStairsF scores for individuals with a given TBI mechanism, and each row shows the scores at the corresponding follow-up year. High-energy transfer TBI mechanisms, which are associated with multisystem trauma (i.e., TBIs from gunshot wounds, assaults, other violence, pedestrian-related accidents, falls, and being struck by falling or flying objects), demonstrate lower median levels of functional stair-climbing, with greater variability over time. Meanwhile, sports-related mechanisms (i.e., TBIs from track and field, gymnastics, water, and air sports) show higher functional levels with minimal variability, often appearing as narrow boxes or near-flat lines in Figure 2. TBIs from gunshot wounds demonstrated the poorest functional recovery trajectory, followed by TBIs from pedestrian-related accidents. The TBI mechanisms were characterized by lower initial median Rasch-transformed FIM Stairs scores and flatter, more stagnant long-term improvement relative to other TBI mechanisms.

Mean Rasch-FIM Stairs scores over 30 years for individuals, grouped by TBI etiology, are shown in Figure 3. Early follow-up years show marked variability in functional stair-climbing. After approximately 10–15 years of follow-up, TBI mechanism recovery trajectories converge, with most TBI mechanisms clustering near moderately negative logit values, suggesting long-term stair-climbing impairments. Most TBI etiologies show either a gradual decline over time or stable plateaus, indicating persistent and ongoing, longstanding difficulties. Differences between mechanism cohorts are most prominent in early follow-up years, suggesting that TBI mechanisms influence initial recovery patterns, whereas long-term outcomes become relatively similar regardless of the TBI mechanism. Vehicular injuries show higher scores during the early follow-up years, followed by sharp declines. The Rasch-FIM Stairs scores for sports-related TBIs show early variability, with pronounced peaks. The scores eventually converge toward negative logit values with other injury mechanisms over time. Individuals with TBIs from falls demonstrate relatively stable recovery trajectories over time. In Rasch terms, the early differences of 0.5–1.0 logits are clinically meaningful changes (57), and the systematic shift toward negative logits in later years suggests long-lasting disability decades after TBI onset, regardless of etiology.

### Significance

The study’s results have paramount clinical and applied significance. By understanding how TBI mechanisms impact very long-term functional outcomes, clinicians can more effectively infer which activities may be more challenging for someone based on their TBI etiology and therefore warrant increased therapeutic attention and practice.

Expediting the procurement of this information enables clinicians to quickly develop and implement TBI mechanism-specific rehabilitation plans. Earlier administration of targeted interventions affords individuals with TBI the opportunity to participate in therapeutic interventions while benefiting from their critical period (or “critical window”) of neuroplasticity, when neurological adaptations enable them to be most responsive to rehabilitation protocols, thereby maximizing their functional improvement following TBI.

The critical period is relatively short-lived. The greatest neuroplastic benefits occur within the first 6 months post-TBI. Subsequently, the beneficial neurological window remains open, but it gradually closes over 6-24 months post-TBI. The time is characterized by a sequence of cellular and molecular events, including axonal sprouting, dendritic remodeling, synaptogenesis, upregulation of NMDA receptor activity, and the downregulation of GABA_A_ receptor activity (58,59).

This study’s findings identify tasks that may be disproportionately difficult for a particular subgroup of individuals with TBI who share a common mechanism of injury. Insights from this study may guide future investigations to refine rehabilitation approaches, optimize functional recovery, and enhance quality of life for individuals living with TBI.

Stair-climbing functional recovery trajectories are illustrated in Figure 2, providing valuable prognostic information and offering individuals with TBI clarity about anticipated recovery patterns. Such knowledge can help determine which actions may likely be managed independently and which may require additional support. Additionally, the results may help inform regional healthcare resource allocation planning. For example, healthcare systems in inner city settings where gun violence may be more prevalent should anticipate an increased demand for neurosurgical and orthopedic services, as well as access to advanced imaging, ventilatory support, and specialized medical equipment. Adequate availability of physical therapy, occupational therapy, and speech-language pathology services is imperative.

Conversely, winter-sports–related TBIs demonstrate higher median logit-FIMStairsF scores with minimal subsequent gains, suggesting that individuals in this group tend to maintain relatively high stair-climbing functionalability throughout their recovery. As such, return-to-work and return-to-school initiatives may be more common in mountainous regions, where TBIs from skiing or snowboarding are more common, necessitating a higher priority for occupational therapy services in these areas. Additionally, demand for assistive technologies, such as wheelchairs, mobility aids, and Augmentative and Alternative Communication (AAC) devices, may exceed the demand in arid desert areas, and the mountainous region may require additional funding and resource allocation to support this population.

### Previous Literature

The results of this study differ from existing literature reporting differences in functional outcomes across TBI mechanisms. This study considered several factors contributing to these differences. First, the sample composition in previous research had often been too homogeneous and biased by regional clustering. This study analyzed a nationally representative cohort with a diverse set of TBI mechanisms and very long-term follow-up, with data collection spanning more than 10 years. Conversely, much of the current literature has relied on smaller samples with shorter follow-up periods that are 10 years or less (often between 2 and 8 years).

Secondly, differences in equipment and outcome measurement methodologies may also contribute to finding discrepancies. Many earlier studies have analyzed raw FIM data, which are susceptible to strong ceiling effects and may skew statistical estimates and standard errors. However, this study transformed the FIMStairsF data using Rasch analysis, yielding FIM logit data, providing an interval-level representation of functional stair-climbing. The transformation ensures that changes are interpreted as equal units of latent functional change across the entire instrument, thereby uncovering group differences that may be missed in analyses using raw FIM data.

Lastly, differences in statistical modeling may yield contradictory results. This study employed a linear mixed-effects modeling framework, which is well-suited to accommodate longitudinal data with unbalanced intervals and allows for the simultaneous estimation of the fixed effects, the TBI mechanism, and the interaction between age and follow-up timing, as well as random effects reflecting sources of individual variability and group clustering. These methodological distinctions explain why this study identified statistically significant and clinically meaningful differences in very long-term functional stair-climbing across TBI mechanisms.

### Strengths & Limitations

Several methodological strengths bolster the rigor and interpretability, enhancing this study’s findings. First, mixed modeling enables the analysis of repeated longitudinal functional outcome measures while accounting for between-group differences and individual-level variability. The methodology is particularly appropriate for longitudinal research, as the models accommodate unequal follow-up intervals and missing data without employing casewise deletion.

Secondly, the FIM Stairs data were converted into interval-level logit values via Rasch analysis before being applied to the mixed models. The transformation accurately reflects latent functional ability and reduces the ceiling effects inherent in raw FIM data. Consequently, comparability across items and time points is optimized, strengthening the validity of longitudinal inferences. Additionally, a strength of this study is the inclusion of multiple TBI mechanisms, which were examined over an extended period, enabling a more detailed characterization of very long-term stair-climbing functional recovery trajectories across TBI mechanisms and yielding clinically meaningful insights that extend well beyond short-term recovery.

Despite the strengths, some limitations also merit consideration. While the sample size is sufficient for mixed modeling, more uncommon TBI mechanisms, such as air sports-related (*n* = 2) and gymnastics-related TBIs (*n* = 1), were represented by relatively small subsamples, reducing statistical power and the generalizability of the study’s results for these groups. Additionally, individuals with long-term follow-up data may differ from those without, potentially introducing selection bias.

While Rasch transformations enhance measurement properties, they also have potential drawbacks: the transformations assume unidimensionality and local independence, and violations of these assumptions may skew interpretations. As for the model structure, the random effects structure may not fully capture all sources of between-person variance. Additionally, confounding variables, such as injury severity, substance abuse, and socioeconomic factors, were not incorporated into the models, which may help explain the observed differences across TBI mechanisms. Lastly, the study modeled time since injury as discrete intervals rather than as a continuous variable. Doing this may obscure nonlinear recovery trajectories.

### Future Research

The results of this study lay a firm foundation for understanding the unique influence of TBI mechanisms on very long-term functional stair-climbing. To build upon these findings, future investigations should address several key methodological limitations. While the TBIMS database provides a comprehensive national sample, more atypical TBI mechanisms, such as gymnastics-related (*n* = 1) and air sports-related (*n* = 2) TBIs, were represented by small subsamples. The limited subsample sizes reduce the statistical power and the generalizability of findings for such TBI mechanisms. Accordingly, future research would benefit from larger sample sizes and multi-site collaborations to support more reliable analyses of less common TBI mechanisms. Additionally, modeling discrete-time post-injury intervals, rather than a continuous-time variable, may not adequately capture the complex, nonlinear patterns of TBI recovery. Exploring nonlinear analytical approaches, such as spline-based growth models, may yield a more nuanced and precise characterization of functional change over time.

Furthermore, for a more comprehensive understanding of TBI recovery, future investigations should extend beyond motor outcomes and should examine cognitive FIM scores. Cognitive functional independence is a primary contributor to one’s quality of life. Investigating the FIM cognitive domain would be a critical next step in determining how TBI mechanisms differentially affect functional independence. Additionally, integrating FIM data with other outcome measures used in TBI rehabilitation, such as the Glasgow Outcome Scale–Extended (GOSE), the Supervision Rating Scale (SRS), and the Participation Assessment with Recombined Tools–Objective (PART-O), would facilitate a more robust, multidimensional evaluation of very long-term community reintegration and recovery trajectories.

Lastly, given the association between TBIs from gunshot wounds and poor very long-term functional outcomes, a closer examination of the disparities is warranted. Incorporating psychosocial factors into future models and investigating the influence of the characteristics on functional outcomes are imperative to advancing this knowledge. Accounting for these factors as potential confounders or mediators in analyses of functional outcomes associated with TBI mechanisms may help clarify the interplay among TBI mechanisms, psychosocial characteristics, and functional recovery, thereby informing and improving the development of targeted, effective, TBI mechanism-specific rehabilitation plans.

## CONCLUSION

This study demonstrates that Traumatic Brain Injury (TBI) mechanisms significantly influence very long-term functional stair-climbing. TBIs from gunshot wounds and pedestrian-related accidents consistently result in poorer functional recovery trajectories, characterized by lower median functional abilities with minimal improvement over time.

The significant negative estimates for TBIs caused by gunshot wounds and pedestrian-related accidents (see Figure 1) suggest that individuals surviving these TBI mechanisms may disproportionately struggle more with their long-term stair-climbing. The findings are clinically significant and enable clinicians to more accurately anticipate the functional challenges their patients may face, based on their TBI mechanism. The insights from this study facilitate the timely development and implementation of mechanism-specific, targeted rehabilitation procedures, enhancing intervention success by leveraging the critical period of neuroplasticity and thereby maximizing the functional improvement of individuals with TBI. The study’s analytical rigor was enhanced by the synergistic application of Rasch analysis, transforming the raw FIMStairsF data into interval-level logit values, mitigating strong ceiling effects and creating a more precise measure of latent stair-climbing functional ability, and linear mixed-effects modeling, which effectively accounted for between-group differences and individual-level variability over longitudinal follow-up periods.

To continue advancing this knowledge, future research should investigate cognitive FIM scores and incorporate psychosocial factors into statistical models, providing a more comprehensive perspective of post-TBI recovery patterns. Developing an understanding of the intricate relationship between TBI mechanisms and very long-term functional outcomes is paramount to creating effective, tailored rehabilitation plans.

## Data Availability

No data available.

